# Brain Imaging-derived Phenotypes and Stroke: A Bidirectional Mendelian Randomization Study Unveils Causal Links between Thalamic Nuclei Volume and Stroke Risk in the European Population

**DOI:** 10.1101/2025.03.22.25324441

**Authors:** Xuanyan Yang, Shiyao Cheng, Shaoli Lin, Peiyue Su, Xinyao Tang, Jiani Wu, Xingchen Lin, Si Cheng, Siyang Liu

## Abstract

**Background and Aims:** Recent advances in brain-imaging techniques have enabled the identification of brain imaging-derived phenotypes (IDPs), representing physiological brain structure. Observational studies have suggested a correlation between these IDPs and stroke, confounded and based on limited samples. To investigate the causal relationship between IDPs and stroke and its subtypes for an in-depth mechanistic comprehension of their interplay, we conducted a bidirectional two-sample Mendelian Randomization (MR) study leveraging the largest-scale genome-wide association studies (GWAS) of IDPs and stroke subtypes.

**Methods:** We utilized GWAS summary statistics from the BIG40 dataset, which included nearly 3,935 IDPs among 33,224 individuals, and GIGASTROKE, which included three etiological ischemia subtypes, as well as cerebral ischemia, intracerebral hemorrhage stroke and overall stroke among 73,652 stroke cases and 1,234,808 controls.

**Results:** In the forward MR analysis, we identified eight significant IDPs influencing the risk of stroke and its subtypes after Bonferroni correction. Notably, the volume of the lateral posterior thalamus in the right hemisphere exhibited a significant negative association with all ischemic stroke (OR=0.79; 95% CI: 0.74 to 0.84; *p*=1.21e-13), all stroke (OR=0.84; 95% CI: 0.79 to 0.89; *p*=2.45e-9), and large vessel stroke (OR=0.54; 95% CI: 0.43 to 0.69; *p*=3.01e-7). Conversely, no significant causal association was observed in the reverse MR analysis.

**Conclusion:** This study enhances our understanding of causality between IDPs and stroke by pinpointing specific causal associations. These findings provide valuable insights into the etiology of stroke, offering potential strategies for predicting and intervening in stroke risk at the level of brain imaging.

## 1. Introduction

Stroke is a prevalent clinical manifestation of cerebrovascular disease and a substantial global public health concern. It encompasses both ischemic and hemorrhagic stroke, characterized by the sudden onset of brain dysfunction(Campbell and Khatri, 2020). The Global Burden of Disease study reported stroke as the second leading cause of death and the third leading cause of death and disability combined in 2019, with ischemic stroke accounting for 62.4% of all cases(Feigin et al., 2021). Although reperfusion therapy has shown efficacy in reducing disability, its practice is constrained by the treatment time window(Campbell et al., 2019). In the absence of effective early prevention strategies, stroke continues to exert a substantial toll on lives and property(Feigin et al., 2021). Consequently, it is crucial to investigate the risk factors contributing to stroke development and devise new preventive strategies.

Brain image-derived phenotypes (IDPs) are digital characteristics of brain tissue obtained through the analysis of brain imaging data, offering precise, reliable, and quantitative information for neuroimaging research(Gong et al., 2021). Advanced imaging techniques enable the prediction of diseases before symptoms manifest, particularly with risk-stratified cohorts(Miller et al., 2016). Numerous observational studies have identified several associations between IDPs and stroke. For instance, periventricular white-matter hyperintensity volume is positively associated with cardioembolic ischemic stroke(Kaffashian et al., 2016). Significant changes in cortical thickness, surface area, and gray matter volume have been observed during stroke recovery(Liu et al., 2023). Patients with cerebral autosomal dominant arteriopathy with subcortical infarcts and leukoencephalopathy exhibit reduced fractional anisotropy (FA), altered mode of anisotropy, and increased mean diffusivity(Zhang et al., 2021). Stroke has also been linked to changes in intra-cellular volume fraction (ICVF), orientation dispersion index (OD), and quantitative T2 values(Siemonsen et al., 2009; Wang et al., 2019). Different IDPs may contribute to stroke subtypes, and conversely, stroke and its subtypes can induce alterations in IDPs. However, existing observational studies often suffer from small sample sizes, and some lack specific detection methods, leaving many aspects of the relationship between stroke and specific brain anatomy or connectivity structures unexplored.

Mendelian randomization (MR) is a statistical approach that utilizes genetic variation as an instrumental variable to assess and quantify causality(Burgess et al., 2016). Genome-wide association studies (GWAS) identify genetic variants associated with disease risk or traits, enabling the exploration of causality through MR. Previous studies have partially unraveled the causal relationship between brain image-derived phenotypes (IDPs) and stroke, with a focus on brain connectivity(Jia et al., 2023; Yu et al., 2023). However, these studies did not investigate the causal link between IDPs related to brain anatomy structures and stroke. Furthermore, these studies have primarily relied on smaller sample sizes derived from the Megastroke study (40,585 cases; 446,696 controls) while the recent Gigastroke has doubled the sample size with 73,652 cases and 1,234,808, controls and unraveled more association signals(Mishra et al., 2022). To obtain the most statistically robust understanding of the causalities between IDPs and stroke risk, we employed a two-sample Mendelian randomization (TSMR) method strategy, complemented by comprehensive sensitive analyses, on the largest scale GWAS summary data available to date.

## 2. Method

### 2.1. Data sources

We employed the summary statistics of brain IDPs from Smith’s GWAS meta-analysis study, which encompassed 3,935 IDPs and 17,103,079 genome-wide SNPs on autosomes 1-22 and the X chromosome. The study comprised a discovery sample of 22,138 individuals and a replication sample of 11,086 individuals of European ancestry(Smith et al., 2021). Detailed data can be accessed on the Oxford Brain Imaging Genetics (BIG40) web server (https://open.win.ox.ac.uk/ukbiobank/big40/). Based on observational studies mentioned before, we selected 10 categories comprising a total of 2,010 IDPs. These categories included 647 regional and tissue volume, 371 cortical area, 303 cortical thickness, 14 regional T2*, 75 white matter (WM) tract FA, 75 WM tract diffusion tensor mode (MO), 300 WM tract diffusivity, 75 WM tract ICVF, 75 WM tract OD, 75 WM tract isotropic or free water volume fraction (ISOVF). **Supplementary Table 2** provides additional information about the data sources for the IDPs.

In our study, we utilized the largest GWAS meta-analysis conducted by the GIGASTROKE consortium to investigate stroke. This analysis included over one million participants, with approximately 70% of European ancestry(Mishra et al., 2022). Comprehensive data can be accessed through the GWAS catalog (https://www.ebi.ac.uk/gwas/). Stroke was categorized into five groups: All Stroke (AS), All Ischemic Stroke (AIS), CardioEmbolic Stroke (CES), Small Vessel Stroke (SVS), and Large Artery Stroke (LAS). The GWAS data in the European population consisted of 1,234,808 controls and varying numbers of cases for each stroke subtype(Mishra et al., 2022). **Supplementary Table 3** provides additional information about the data sources for stroke and its subtypes.

### 2.2 Study design

To investigate the causality between IDPs and stroke, we followed a four-step process based on Burgess’s study for Bidirectional Two-Sample Mendelian Randomization(Burgess et al., 2016). Firstly, we collected GWAS summary data from public studies or websites mentioned earlier and organized the data into categories. Next, we selected SNPs as instrumental variables (IVs) for each IDP and stroke type based on three IV hypotheses. Thirdly, we conducted a bidirectional two-sample MR using five methods separately for the causal directions of IDPs to stroke and stroke to IDPs. Lastly, we performed a sensitivity analysis to validate the IVs and ensure the reliability of the MR results. The workflow and a brief overview are presented in **Fig. 1**.

**Fig. 1.**
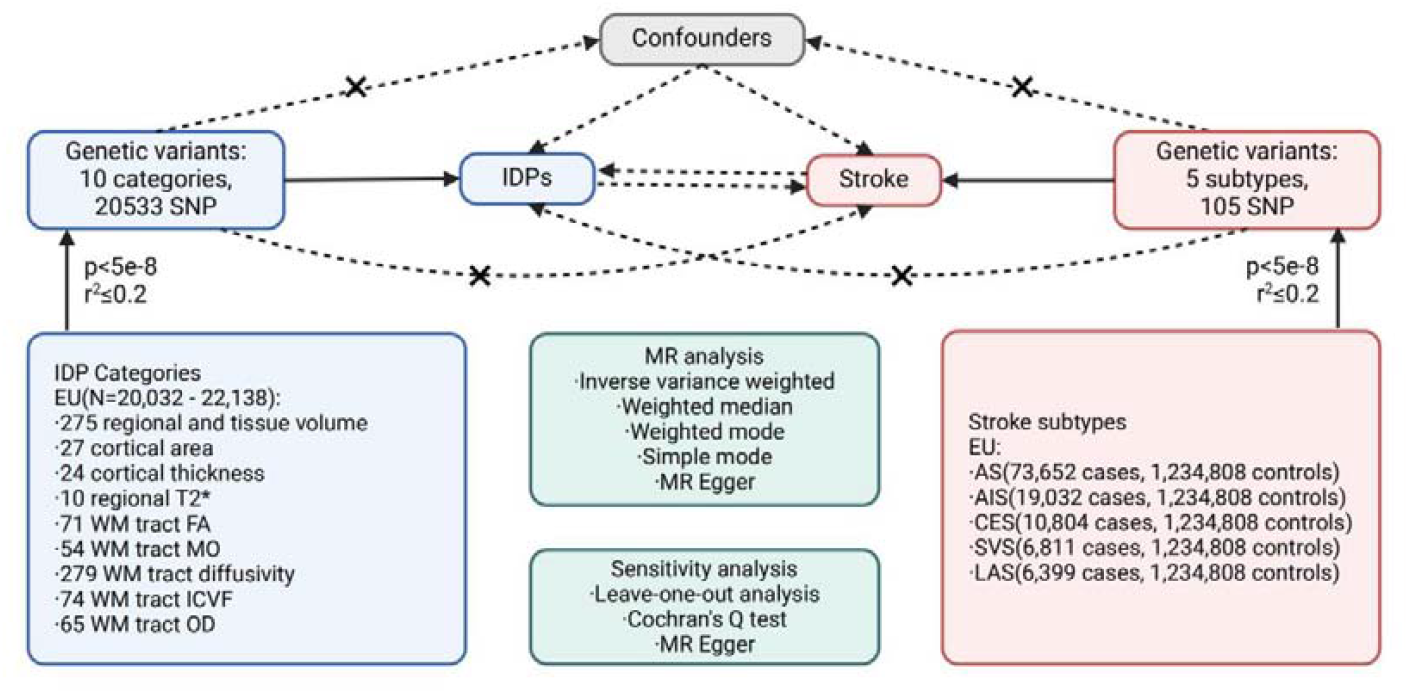
Workflow of the causal inference between IDPs and stroke.

### 2.3. Instrumental variables selection

The three IV assumptions, as outlined by MR guidelines(Burgess et al., 2016), are as follows: 1) The variants exhibit a strong association with the exposure, 2) the variants are not associated with the outcome through any confounding factors, 3) the variants do not directly impact the outcome, except via the exposure-outcome pathway. To select independent SNPs as IVs, we employed genome-wide conditional & joint association analysis (GCTA-COJO)(Yang et al., 2011). This approach evaluates the variance explained by all SNPs on a chromosome or genome for a specific disease or trait to ensure the independence of multiple variants at a given locus. We applied a p-value threshold of 5e-8 and a linkage disequilibrium r-square less than 0.2 (gcta64 –bfile reference_panel_file –cojo-file gwas_file –cojo—slct –cojo-p 5e-8 --cojo-collinear 0.2) to select satisfactory SNPs, which were then included as IVs in our subsequent study. **Supplementary Table 2** provides further information on the IVs for IDPs, while **Supplementary Table 3** contains information on the IVs for stroke. The source code and data are openly accessible in online Github repository (https://github.com/liusylab/IDP_Stroke_2SMR).

### 2.4. Statistical analysis

The MR analysis process details are outlined in the STROBE checklist. At first, we used the harmonise_data() function from the TwoSampleMR package to harmonize the exposure and outcome data. Then, we conducted bidirectional Two Sample Mendelian Randomization (TSMR) using five methods, including inverse variance weighted (IVW), MR Egger, weighted mode, simple mode, and weighted median. IVW was chosen as the primary method to explore the causal relationships between IDPs and stroke due to its robustness(Hartwig et al., 2017), while the other four methods were used to further validate the results. Sensitivity analysis was performed using MR Egger to assess horizontal pleiotropy(Bowden et al., 2015), and leave-one-out analysis and Cochran’s Q test were also conducted. Leave-one-out analysis evaluated the reliability of the MR model, visually represented by a forest plot. Cochran’s Q test was employed to identify directional heterogeneity among SNPs. Additionally, MR Steiger analysis was performed to estimate the sensitivity ratio and assess the correctness of the causal direction. All statistical analyses were conducted using the TwoSampleMR packages in R version 4.2.2. To account for multiple testing, we applied Bonferroni correction across the entire MR analysis, setting the significance threshold at *p*<2.46e-6 (0.05/ (2010*5*2)), where 2010*5 represents the number of all IDP-stroke pairs and 2 denotes forward and reverse MR tests.

## 3. Results

In the forward MR analysis, a total of 6,363 TSMR tests were conducted to assess the causal relationship between 1,289 IDPs and 5 stroke types. Using the robust IVW method, 733 results reached nominal significance (*p*<0.05), involving 517 IDPs (**Fig. 2, Supplementary Table 4**). Following the Bonferroni correction for multiple testing, we identified eight significant causal relationships between four IDPs and three stroke types. These associations were all within the same brain anatomy categories and specifically related to the volume of thalamus nuclei. All the results from the five MR methods, under the multiple testing correction, are provided in **Fig. 3** and visualized with the location and structure of the thalamus in **Fig. 4**.

**Fig. 2.**
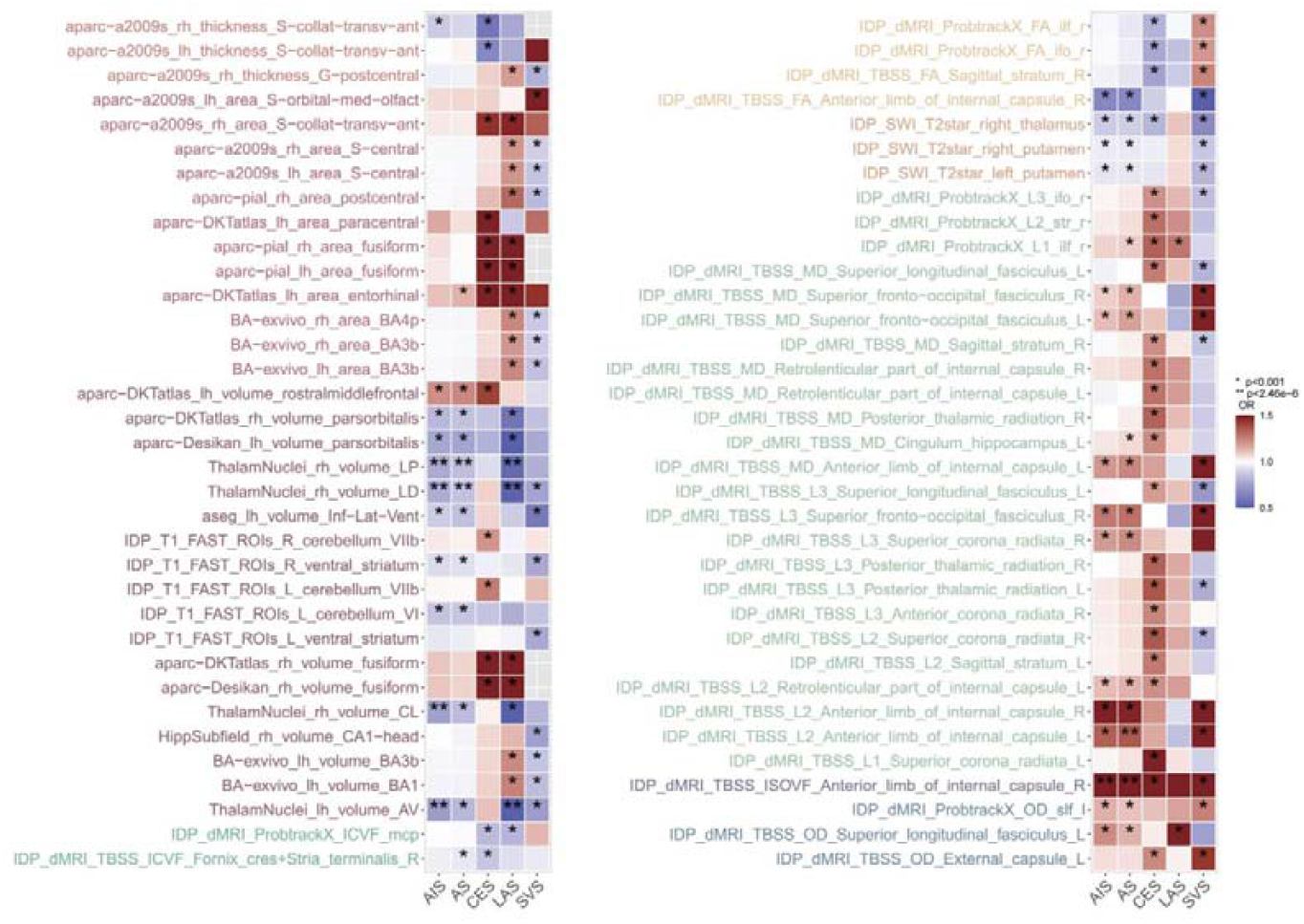
Overview of the forward MR based on IVW. (**A)** the result for IDPs to the stroke (regional and tissue volume, cortical area, cortical thickness, WM tract ICVF), (**B)** the result for stroke to IDPs (regional T2*, WM tract FA, WM tract diffusivity, WM tract OD, WM tract ISOVF). The p-value threshold was set as 0.001 (0.05/ (10*5), 10 donates a number of IDP categories, and 5 donates a number of stroke types). Only associations that existed in the relationship between IDPs and stroke are listed in **Fig. 2**.

**Fig. 3.**
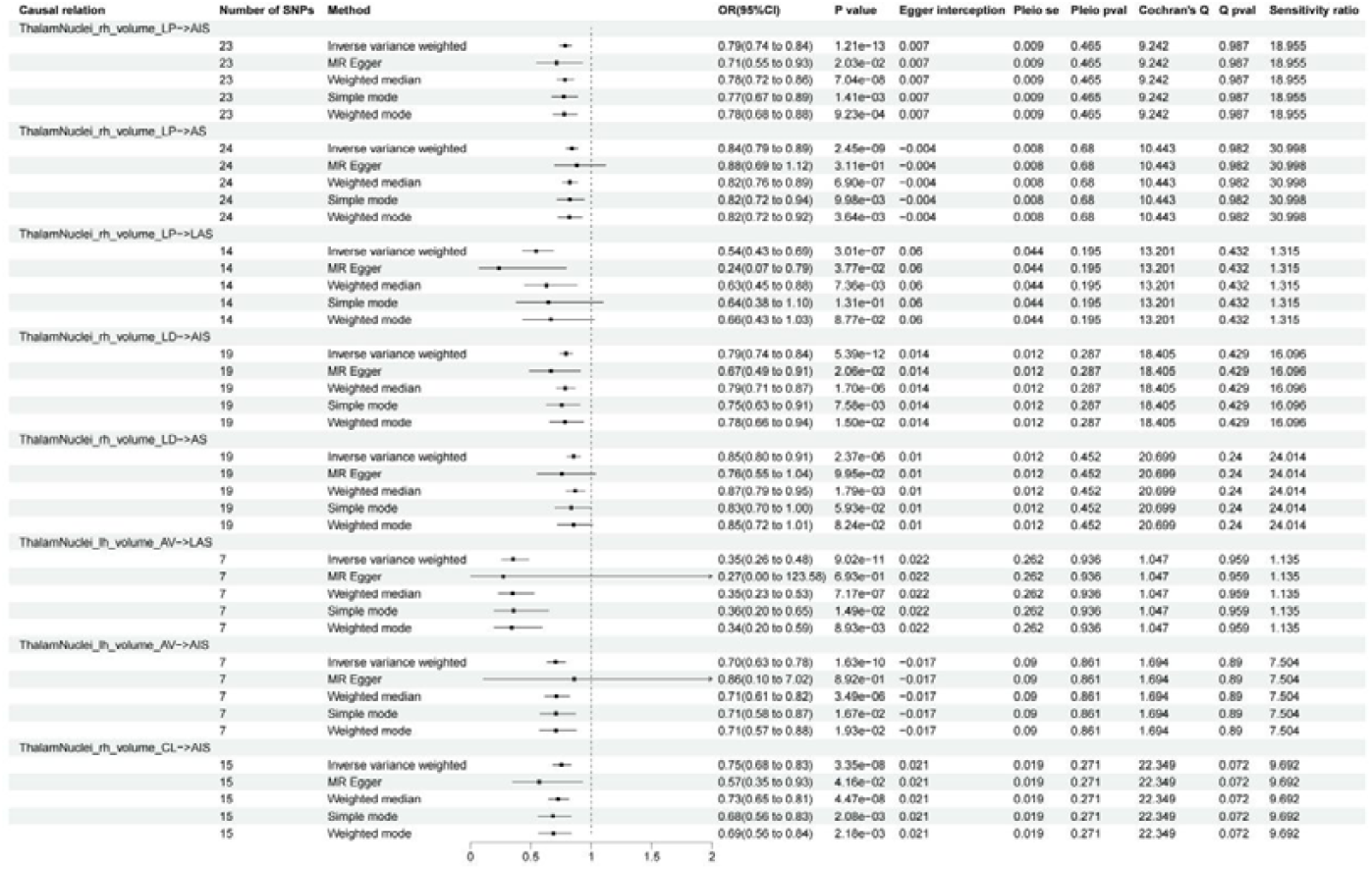
Causalities in the forward MR. Only results that remain significant after Bonferroni correction are listed (*p*<2.46e-6). The arrow implies the maximum interval extended on the axis.

**Fig. 4.**
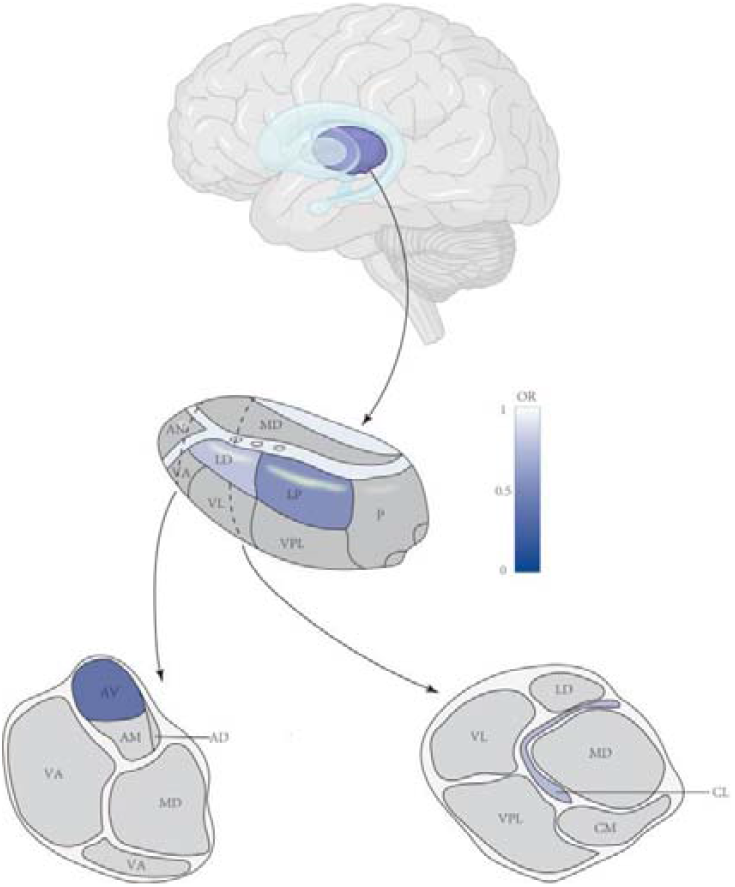
Schematic diagram of thalamic nuclei. Abbreviations: AN, anterior nucleus; LD, lateral dorsal; LP, lateral posterior; MD, medial dorsal; VA, ventral anterior; VL, ventral lateral; VPL, ventral posterior lateral; P, pulvinar; AV, anterior ventral;AM, anterior medial; AD, anterior dorsal; CL, central lateral; CM, central medial. The thalamus nuclei showed no association with stroke and painted gray. The shades of LP, LD, AV, and CL are linked with minimum OR.

The lateral posterior nucleus of the thalamus (LP) receives extensive afferent fibers from the cerebral cortex. An increase of 1 s.d. in the volume of LP in the right hemisphere was associated with a 21% lower risk of AIS (OR=0.79; 95% CI: 0.74 to 0.84; *p*=1.21e−13), a 16% lower risk of AS (OR=0.84; 95% CI: 0.79 to 0.89; *p*=2.45e −9), and a 46% lower risk of LAS (OR=0.54; 95% CI: 0.43 to 0.69; *p*=3.01e−7). The lateral dorsal nucleus of the thalamus (LD) showed that an increase of 1 s.d. in its volume in the right hemisphere was associated with a 21% lower risk of AIS OR=0.79; 95% CI: 0.74 to 0.84; *p*=5.39e−12) and a 15% lower risk of AS (OR=0.85; 95% CI: 0.80 to 0.91; *p*=2.37e−6). The anterior ventral nucleus of the thalamus (AV) demonstrated that an increase of 1 s.d. in its volume in the left hemisphere was associated with a 65% lower risk of LAS (OR=0.35; 95% CI: 0.26 to 0.48; *p*=9.02e− 11) and a 30% lower risk of AIS (OR=0.70; 95% CI: 0.63 to 0.78; *p*=1.63e-10). Similarly, the central lateral nucleus of the thalamus (CL) displayed that an increase of 1 s.d. in its volume in the right hemisphere was associated with a 25% lower risk of AIS (OR=0.75; 95% CI: 0.68 to 0.83; *p*=3.35e−8). Four significant IDPs are causally linked to AIS, with corresponding scatter plots presented in **Fig. 5**. Additional scatter plots for other pairs can be found in **Supplementary Figure 2-5**.

**Fig. 5.**
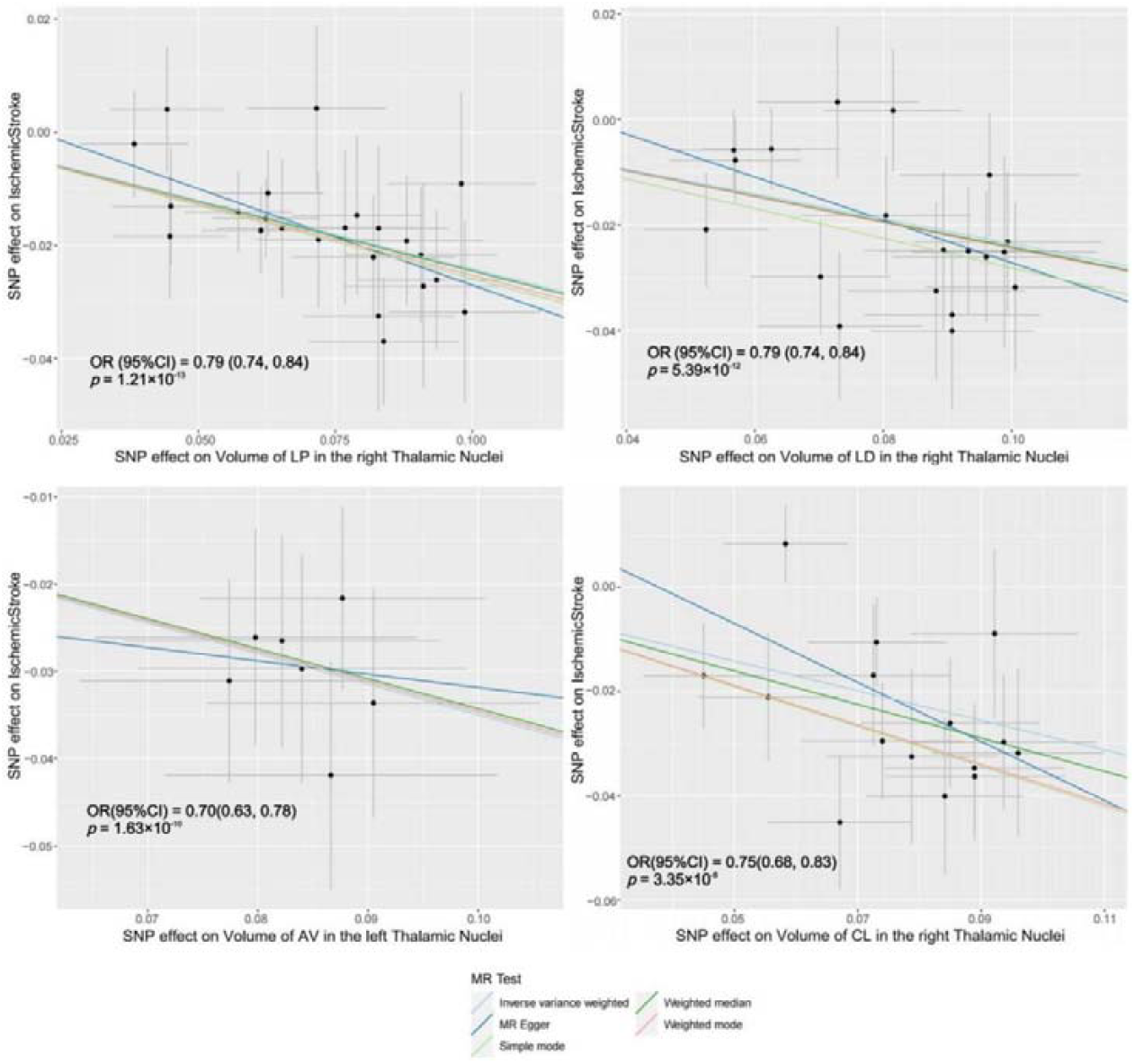
Scatter plots of individual SNP effects and estimates from different MR methods. **(A)** Volume of LP in the right Thalamic Nuclei to Ischemic Stroke, (**B)** Volume of LD in the right Thalamic Nuclei to Ischemic Stroke, (**C)** Volume of AV in the left Thalamic Nuclei to Ischemic Stroke, (**D)** Volume of CL in the left Thalamic Nuclei to Ischemic Stroke.

Horizontal pleiotropy was assessed using the intercept term of MR Egger regression and funnel plots, which showed no evidence of its presence (**Fig. 3, Supplementary Figure 6-13**). Directional heterogeneity was evaluated through Cochran’s Q test, which revealed no evidence of such heterogeneity (**Fig. 3**). The leave-one-out analysis demonstrated minimal change in the results after removing any individual SNP, indicating the high reliability of the MR models (**Supplementary Figure 14-21**). In summary, the sensitivity analyses confirmed the reliability of our forward MR results.

In the reverse MR analysis, we performed 8,241 tests to examine the causal relationship between stroke and IDPs (**Supplementary Table 5**). Using IVW, we identified 554 nominal significant results (*p*<0.05), involving 464 IDPs (**Supplementary Figure 1**). However, after applying the Bonferroni correction, none of the results remained significant.

## 4. Discussion

Our study conducted bidirectional TSMR analyses to comprehensively investigate the causal relationship between brain IDPs and stroke or its subtypes in the European population. To ensure statistical validity, we utilized the largest available cohorts with genetic information for IDPs and stroke. We identified several IDPs that showed nominal significance with stroke and its subtypes in both causal directions (p<0.05). After correcting for multiple tests, eight associations remained significant in the forward MR analysis, while no results remained significant in the reverse MR analysis.

In the forward MR analysis, we identified four IDPs causally associated with three stroke types, all related to the volume of thalamus nuclei. LP, a thalamus nucleus involved in visual processing(Casanova et al., 1991) and rotation response(Motles et al., 1985), showed negative associations with AIS, AS, and LAS. In LP, stroke manifestations commonly include hemihypesthesia, hemiataxia, and executive dysfunction, typically resulting from artery embolism and microangiopathy(Carrera et al., 2004). However, there is currently no observational evidence linking the volume of LP and stroke. LD, a thalamus nucleus connected to the posterior parietal cortex and cingulate gyrus(Shibata and Naito, 2005), which play roles in spatial learning and memory processing(Bezdudnaya and Keller, 2008), demonstrated negative associations with AIS and AS. Unfortunately, no relevant studies have explored the relationship between the volume of LD and stroke. AV, a thalamus nucleus with connections to the frontal cortex(Yeterian and Pandya, 1988) and cingulate gyrus(Baleydier and Mauguiere, 1980), linked to behavioral activation(c-Fos expression in the limbic thalamus following thermoregulatory and wake-sleep changes in the rat - PubMed, n.d.) and learning functions(Shibata and Yoshiko, 2015), exhibited a negative correlation between the volume of the left AV and LAS risk.

Although the volume of AV has been reported to decrease in psychiatric disorders like schizophrenia(Forno et al., 2023), no similar study has investigated the association between AV and stroke. CL, a thalamus nucleus forming part of the rostral intralaminar nuclei, receiving inputs from limbic, sensorimotor-related, and cerebellar input, and distributing over the frontal cortex and dorsal striatum(Vertes et al., 2022), has potential functions in arousal(Xu et al., 2020) and motor control(Sakayori et al., 2019). Our results indicated that a decrease in the volume of the right CL was associated with a higher risk of AIS. However, no studies have measured the volume of CL or explored its relationship with stroke. Previous research suggests that white matter loss after thalamic infarction may derive the cortical reshaping(Conrad et al., 2022). Stroke can be caused by cerebral vessels with different anatomical structures through various mechanisms(Kim et al., 2019). Based on these findings, we hypothesize that a genetically predicted smaller thalamic nucleus volume may result in more tortuous nerve fibers and blood vessels, increasing the risk of stroke.

While the evidence regarding specific thalamus nuclei and stroke types is limited, the thalamus has been consistently associated with stroke. Children with perinatal AIS exhibit a smaller volumes of both thalami compared to control groups(Ilves et al., 2022). Similarly, non-thalamic stroke patients also display smaller thalamic volume on both sides(Geng et al., 2023). Most observational studies have reported smaller volumes in the ipsilateral or bilateral thalamus, which may partially align with our findings but requires further investigation for confirmation. Assuming that the subcortical grey matter remains similar before and after a stroke within six weeks(Predicted Brain Age After Stroke - PubMed, n.d.), the smaller thalamic volume before the stroke likely has an adverse impact by limiting resources(Liew et al., 2021).

Though clinical and observational studies have indicated that stroke can lead to various changes in IDPs, we did not establish a causal relationship between stroke and IDPs in our reverse MR analyses. Since the imaging data was obtained from a healthy population, it is expected not to observe any reverse causality. On the other hand, this may suggest that the intricate mechanisms of neural connections account for the absence of causality, or the effects are too subtle to detect in our study.

Our findings differ significantly from previous bidirectional TSMR findings by Yu et al., published in BMC Medicine(Yu et al., 2023), and Jia et al., in Cerebral Cortex(Jia et al., 2023). Since Jia’s work is similar to that of BMC, but with a narrower range, we primarily compare and discuss our results with those from Yu. Yu identified potential causal effects between decreased FA, increased MD, and ISOVF on stroke in the forward MR analysis, as well as the causal effect of stroke on ISOVF in the reverse MR analysis. However, these associations were not observed in our study. A comparison of the two studies is presented in **Supplementary Table 6**. The disparity in results could be attributed to two main reasons. Firstly, we utilized the largest stroke GWAS results from GIGASTROKE(Mishra et al., 2022), while Yu’ s study used data from MEGASTROKE(Yu et al., 2023). The utilization of a larger GWAS dataset enhances the power to assess causal relationships while reducing false positives. Secondly, the screening criteria for selecting IVs differed between the two studies, which may have resulted in variations in the IVs chosen. We were unable to replicate Yu’s tests using their IVs due to errors in their provided repetitive IV information. Additionally, the significant IDP-stroke pairs identified in our study were not included in Yu’s study due to their artificial selection of the IDPs before the MR study.

Our study employed bidirectional TSMR using SNPs as instrumental variables to investigate the causal relationships between IDPs and stroke. This approach mitigated environmental or lifestyle confounding factors and demonstrated greater effectiveness compared to observational case-control studies. Additionally, we utilized the largest available GWAS summary data for both exposure and outcome, enhancing statistical power. Our findings revealed a potential causal association between the volume of thalamus nuclei and the development of stroke, a previously unexplored relationship.

Despite the advantages, there are a few limitations of our study. The selection of traits for bidirectional TSMR analysis was not based on careful consideration of existing observational studies, highlighting the need for further research to determine the exact nature of these relationships. However, screening of the IDPs without selection of the traits also ensures a comprehensive understanding of the causality relationships between IDPs and stroke. The population under investigation in this study is limited to individuals of European descent due to data availability constraints. Nevertheless, the insights gained and the methodologies employed in this research can serve as a valuable reference for future investigations among non-European populations.

Additionally, we applied a conservative multiple-testing correction, which may have led to the exclusion of previously reported or potentially significant associations, but it guaranteed the robustness of our results. For instance, our study did not find a significant causal association between MD in the superior fronto-occipital fasciculus and stroke, despite its reported significance in Yu’s work(Yu et al., 2023). This discrepancy could be attributed to differences in IV screening conditions and/or the utilization of a larger database. Further validation on which is the reasons for the discrepancy will require a complete release of the IVs in Yu’s work. We have made our IVs completely publicly available to facilitate the reproducibility of our study.

## 5. Conclusion

In summary, our study utilized bidirectional TSMR analysis with the largest available GWAS summary data to investigate the causal relationship between brain IDPs and stroke or its subtypes. We identified causal associations between the volume of thalamus nuclei and stroke, contributing to our understanding of the link between brain anatomy structure and stroke. These findings may have implications for potential strategies in predicting and intervening in stroke risk at the brain imaging level. By identifying individuals with specific brain IDPs that potentially lead to stroke, clinicians can recommend appropriate lifestyle changes or monitor their health status accordingly. This could facilitate the implementation of high-risk strategies more conveniently.

## Data Availability

All data produced in the present study are available upon reasonable request to the authors.
All data produced in the present work are contained in the manuscript.
All data produced are available online at https://github.com/liusylab/IDP_Stroke_2SMR.

## Abbreviations

IDP: Imaging-Derived Phenotype
MR: Mendelian Randomization
GWAS: Genome-Wide Association Study
AS: All Stroke
AIS: All Ischemic Stroke
LAS: Large Vessel Stroke
CES: CardioEmbolic Stroke
SVS: Small Vessel Stroke
FA: fractional anisotropy
ICVF: Intra-Cellular Volume Fraction
OD: Orientation Dispersion index
TSMR: two-sample Mendelian randomization
ISOVF: Isotropic or free water Volume Fraction
IV: Instrumental Variables
IVW: Inverse Variance Weighted
LP: Lateral Posterior nucleus of the thalamus
LD: Lateral Dorsal nucleus of the thalamus
AV: Anterior Ventral nucleus of the thalamus
CL: Central Lateral nucleus of the thalamus

## Author contributions

The study was conceived and designed by SL, SYC. GWAS summary data was collected by SYC, XCL, XYY, LSL, PYS, YXT, JNW. IV selection, MR analysis and manuscript were done by XYY. Manuscript was revised by SL, SYC, XYY.

## Ethics approval statement

The study has obtained ethical approval as documented in previous GWAS research. Participants have provided informed consent prior to their participation in the study.

## Data availability

The GWAS summary statistics utilized in our study were sourced from the largest scale published GWA studies on both IDPs and stroke. Further information of our study are provided in **Supplementary tables** and **Supplementary Figures**. All supplementary data for this study are accessible in online open-access repository (https://github.com/liusylab/IDP_Stroke_2SMR).

## Appendix A. Supplementary Tables

Further information about the data we used can be obtained in **Supplementary Tables**.

## Appendix B. Supplementary Figures

Further illustrations of our study is presented in **Supplementary Figures**.

## Acknowledgement

This study is supported by grants from National Key R&D Program of China (2022YFC2502400, 2022YFC2502402, 2022YFE0209600), the National Natural Science Foundation of China (82101359), Young Elite Scientists Sponsorship Program by CAST (2023QNRC001).

## Reference

Baleydier, C., and Mauguiere, F. (1980). The duality of the cingulate gyrus in monkey. Neuroanatomical study and functional hypothesis. Brain 103, 525–554. doi: 10.1093/brain/103.3.525

Bezdudnaya, T., and Keller, A. (2008). Laterodorsal nucleus of the thalamus: A processor of somatosensory inputs. J Comp Neurol 507, 1979–1989. doi: 10.1002/cne.21664

Bowden, J., Davey Smith, G., and Burgess, S. (2015). Mendelian randomization with invalid instruments: effect estimation and bias detection through Egger regression. Int J Epidemiol 44, 512–525. doi: 10.1093/ije/dyv080

Burgess, S., Dudbridge, F., and Thompson, S. G. (2016). Combining information on multiple instrumental variables in Mendelian randomization: comparison of allele score and summarized data methods. Stat Med 35, 1880–1906. doi: 10.1002/sim.6835

Campbell, B. C. V., De Silva, D. A., Macleod, M. R., Coutts, S. B., Schwamm, L. H., Davis, S. M., et al. (2019). Ischaemic stroke. Nat Rev Dis Primers 5, 70. doi: 10.1038/s41572-019-0118-8

Campbell, B. C. V., and Khatri, P. (2020). Stroke. Lancet 396, 129–142. doi: 10.1016/S0140-6736(20)31179-X

Carrera, E., Michel, P., and Bogousslavsky, J. (2004). Anteromedian, central, and posterolateral infarcts of the thalamus: three variant types. Stroke 35, 2826–2831. doi: 10.1161/01.STR.0000147039.49252.2f

Casanova, C., Nordmann, J. P., and Molotchnikoff, S. (1991). [Pulvina-lateralis posterior nucleus complex of mammals and the visual function]. J Physiol (Paris) 85, 44–57.

c-Fos expression in the limbic thalamus following thermoregulatory and wake-sleep changes in the rat - PubMed (n.d.). Available at: https://pubmed.ncbi.nlm.nih.gov/30887077/ (Accessed March 3, 2024).

Conrad, J., Habs, M., Ruehl, R. M., Bögle, R., Ertl, M., Kirsch, V., et al. (2022). White matter volume loss drives cortical reshaping after thalamic infarcts. Neuroimage Clin 33, 102953. doi: 10.1016/j.nicl.2022.102953

Feigin, V. L., Stark, B. A., Johnson, C. O., Roth, G. A., Bisignano, C., Abady, G. G., et al. (2021). Global, regional, and national burden of stroke and its risk factors, 1990–2019:a systematic analysis for the Global Burden of Disease Study 2019. The Lancet Neurology 20, 795–820. doi: 10.1016/S1474-4422(21)00252-0

Forno, G., Saranathan, M., Contador, J., Guillen, N., Falgàs, N., Tort-Merino, A., et al. (2023). Thalamic nuclei changes in early and late onset Alzheimer’s disease. Curr Res Neurobiol 4, 100084. doi: 10.1016/j.crneur.2023.100084

Geng, J., Gao, F., Ramirez, J., Honjo, K., Holmes, M. F., Adamo, S., et al. (2023). Secondary thalamic atrophy related to brain infarction may contribute to post-stroke cognitive impairment. J Stroke Cerebrovasc Dis 32, 106895. doi: 10.1016/j.jstrokecerebrovasdis.2022.106895

Gong, W., Beckmann, C. F., and Smith, S. M. (2021). Phenotype discovery from population brain imaging. Med Image Anal 71, 102050. doi: 10.1016/j.media.2021.102050

Hartwig, F. P., Davey Smith, G., and Bowden, J. (2017). Robust inference in summary data Mendelian randomization via the zero modal pleiotropy assumption. Int J Epidemiol 46, 1985–1998. doi: 10.1093/ije/dyx102

Ilves, N., Lõo, S., Ilves, N., Laugesaar, R., Loorits, D., Kool, P., et al. (2022). Ipsilesional volume loss of basal ganglia and thalamus is associated with poor hand function after ischemic perinatal stroke. BMC Neurol 22, 23. doi: 10.1186/s12883-022-02550-3

Jia, Y., Sun, H., Sun, L., Wang, Y., Xu, Q., Liu, Y., et al. (2023). Mendelian randomization analysis implicates bidirectional associations between brain imaging-derived phenotypes and ischemic stroke. Cereb Cortex 33, 10848–10857. doi: 10.1093/cercor/bhad329

Kaffashian, S., Tzourio, C., Zhu, Y.-C., Mazoyer, B., and Debette, S. (2016). Differential Effect of White-Matter Lesions and Covert Brain Infarcts on the Risk of Ischemic Stroke and Intracerebral Hemorrhage. Stroke 47, 1923–1925. doi: 10.1161/STROKEAHA.116.012734

Kim, Y. S., Kim, B. J., Noh, K. C., Lee, K. M., Heo, S. H., Choi, H.-Y., et al. (2019). Distal versus Proximal Middle Cerebral Artery Occlusion: Different Mechanisms. Cerebrovasc Dis 47, 238–244. doi: 10.1159/000500947

Liew, S.-L., Zavaliangos-Petropulu, A., Schweighofer, N., Jahanshad, N., Lang, C. E., Lohse, K. R., et al. (2021). Smaller spared subcortical nuclei are associated with worse post-stroke sensorimotor outcomes in 28 cohorts worldwide. Brain Commun 3, fcab254. doi: 10.1093/braincomms/fcab254

Liu, J., Wang, C., Qin, W., Ding, H., Peng, Y., Guo, J., et al. (2023). Cortical structural changes after subcortical stroke: Patterns and correlates. Hum Brain Mapp 44, 727–743. doi: 10.1002/hbm.26095

Miller, K. L., Alfaro-Almagro, F., Bangerter, N. K., Thomas, D. L., Yacoub, E., Xu, J., et al. (2016). Multimodal population brain imaging in the UK Biobank prospective epidemiological study. Nat Neurosci 19, 1523–1536. doi: 10.1038/nn.4393

Mishra, A., Malik, R., Hachiya, T., Jürgenson, T., Namba, S., Posner, D. C., et al. (2022). Stroke genetics informs drug discovery and risk prediction across ancestries. Nature 611, 115–123. doi: 10.1038/s41586-022-05165-3

Motles, E., Infante, C., and González, M. (1985). [Role of the pulvinar lateralis posterior complex on the rotation response. Relation with other cerebral structures. Pharmacological systems involved in this behavior]. Acta Physiol Pharmacol Latinoam 35, 237–249.

Predicted Brain Age After Stroke - PubMed (n.d.). Available at: https://pubmed.ncbi.nlm.nih.gov/31920628/ (Accessed March 20, 2024).

Sakayori, N., Kato, S., Sugawara, M., Setogawa, S., Fukushima, H., Ishikawa, R., et al. (2019). Motor skills mediated through cerebellothalamic tracts projecting to the central lateral nucleus. Mol Brain 12, 13. doi: 10.1186/s13041-019-0431-x

Shibata, H., and Naito, J. (2005). Organization of anterior cingulate and frontal cortical projections to the anterior and laterodorsal thalamic nuclei in the rat. Brain Res 1059, 93–103. doi: 10.1016/j.brainres.2005.08.025

Shibata, H., and Yoshiko, H. (2015). Thalamocortical projections of the anteroventral thalamic nucleus in the rabbit. J Comp Neurol 523, 726–741. doi: 10.1002/cne.23700

Siemonsen, S., Mouridsen, K., Holst, B., Ries, T., Finsterbusch, J., Thomalla, G., et al. (2009). Quantitative t2 values predict time from symptom onset in acute stroke patients. Stroke 40, 1612–1616. doi: 10.1161/STROKEAHA.108.542548

Smith, S. M., Douaud, G., Chen, W., Hanayik, T., Alfaro-Almagro, F., Sharp, K., et al. (2021). An expanded set of genome-wide association studies of brain imaging phenotypes in UK Biobank. Nat Neurosci 24, 737–745. doi: 10.1038/s41593-021-00826-4

Vertes, R. P., Linley, S. B., and Rojas, A. K. P. (2022). Structural and functional organization of the midline and intralaminar nuclei of the thalamus. Front Behav Neurosci 16, 964644. doi: 10.3389/fnbeh.2022.964644

Wang, Z., Zhang, S., Liu, C., Yao, Y., Shi, J., Zhang, J., et al. (2019). A study of neurite orientation dispersion and density imaging in ischemic stroke. Magnetic Resonance Imaging 57, 28–33. doi: 10.1016/j.mri.2018.10.018

Xu, J., Galardi, M. M., Pok, B., Patel, K. K., Zhao, C. W., Andrews, J. P., et al. (2020). Thalamic Stimulation Improves Postictal Cortical Arousal and Behavior. J Neurosci 40, 7343–7354. doi: 10.1523/JNEUROSCI.1370-20.2020

Yang, J., Lee, S. H., Goddard, M. E., and Visscher, P. M. (2011). GCTA: a tool for genome-wide complex trait analysis. Am J Hum Genet 88, 76–82. doi: 10.1016/j.ajhg.2010.11.011

Yeterian, E. H., and Pandya, D. N. (1988). Corticothalamic connections of paralimbic regions in the rhesus monkey. J Comp Neurol 269, 130–146. doi: 10.1002/cne.902690111

Yu, K., Chen, X.-F., Guo, J., Wang, S., Huang, X.-T., Guo, Y., et al. (2023). Assessment of bidirectional relationships between brain imaging-derived phenotypes and stroke: a Mendelian randomization study. BMC Med 21, 271. doi: 10.1186/s12916-023-02982-9

Zhang, Q., Wang, D., Wu, S., Ren, Y., Li, Y., Zhang, J., et al. (2021). Diffuse Tract Damage Correlates With Global Cognitive Impairment in Cerebral Autosomal Dominant Arteriopathy With Subcortical Infarcts and Leukoencephalopathy: A Tract-Based Spatial Statistics Study. J Comput Assist Tomogr 45, 285–293. doi: 10.1097/RCT.0000000000001129

