## Supplementary Figures for "Brain Imaging-derived Phenotypes and Stroke: A Bidirectional Mendelian Randomization Study Unveils Causal Links between Thalamic Nuclei Volume and Stroke Risk in the European Population"

This supplementary material is provided by authors to give additional information about our works. **Supplementary Figure 1** is the overview of the causality between stroke and IDPs. **Supplementary Figure 2** to **5** are scatter plots. **Supplementary Figure 6** to **13** are funnel plots. **Supplementary Figure 14** to **21** are results of MR leave-one-out sensitivity analysis.

**Supplementary Figure 1** The overview of the causality between stroke and IDPs

**Supplementary Figure 2** Scatter plot of individual SNP effects and estimates from different MR methods, Volume of LP in the right Thalamic Nuclei to Stroke

**Supplementary Figure 3** Scatter plot of individual SNP effects and estimates from different MR methods, Volume of LP in the right Thalamic Nuclei to Large Artery Stroke

**Supplementary Figure 4** Scatter plot of individual SNP effects and estimates from different MR methods, Volume of AV in the left Thalamic Nuclei to Large Artery Stroke

**Supplementary Figure 5** Scatter plot of individual SNP effects and estimates from different MR methods, Volume of LD in the right Thalamic Nuclei to Stroke

**Supplementary Figure 6** Funnel plot of Volume of LP in the right Thalamic Nuclei to Ischemic Stroke

**Supplementary Figure 7** Funnel plot of Volume of LP in the right Thalamic Nuclei to Stroke

**Supplementary Figure 8** Funnel plot of Volume of LP in the right Thalamic Nuclei to Large Artery Stroke

**Supplementary Figure 9** Funnel plot of Volume of AV in the left Thalamic Nuclei to Large Artery Stroke

**Supplementary Figure 10** Funnel plot of Volume of AV in the left Thalamic Nuclei to Ischemic Stroke

**Supplementary Figure 11** Funnel plot of Volume of CL in the right Thalamic Nuclei to Ischemic Stroke

**Supplementary Figure 12** Funnel plot of Volume of LD in the right Thalamic Nuclei to Ischemic Stroke

**Supplementary Figure 13** Funnel plot of Volume of LD in the right Thalamic Nuclei to Stroke

**Supplementary Figure 14** MR leave-one-out sensitivity analysis of Volume of LP in the right

Thalamic Nuclei to Ischemic Stroke

**Supplementary Figure 15** MR leave-one-out sensitivity analysis of Volume of LP in the right Thalamic Nuclei to Stroke

**Supplementary Figure 16** MR leave-one-out sensitivity analysis of Volume of LP in the right Thalamic Nuclei to Large Artery Stroke

**Supplementary Figure 17** MR leave-one-out sensitivity analysis of Volume of AV in the left Thalamic Nuclei to Large Artery Stroke

**Supplementary Figure 18** MR leave-one-out sensitivity analysis of Volume of AV in the left Thalamic Nuclei to Ischemic Stroke

**Supplementary Figure 19** MR leave-one-out sensitivity analysis of Volume of CL in the right Thalamic Nuclei to Ischemic Stroke

**Supplementary Figure 20** MR leave-one-out sensitivity analysis of Volume of LD in the right Thalamic Nuclei to Ischemic Stroke

**Supplementary Figure 21** MR leave-one-out sensitivity analysis of Volume of LD in the right Thalamic Nuclei to Stroke

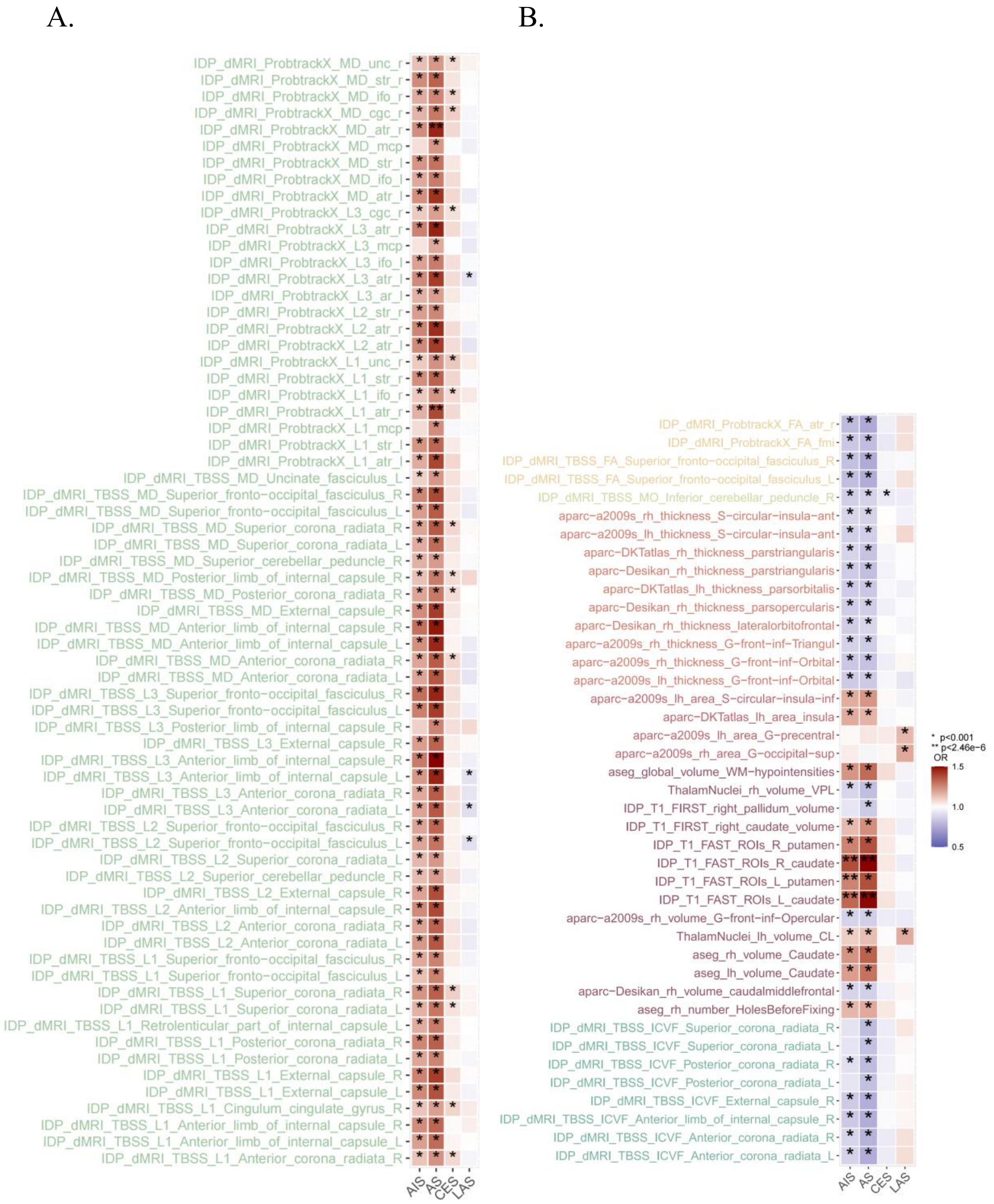


**Supplementary Figure 1** The overview of the causality between stroke and IDPs, the results are based on IVW method. **A** the result for stroke to the IDPs (WM tract diffusivity), **B** the result for stroke to IDPs (regional and tissue volume, cortical area, cortical thickness, WM tract FA, WM tract ICVF, WM tract MO). Only associations existed in the relationship between stroke and IDPs are listed in the **Supplementary Figure 1**.

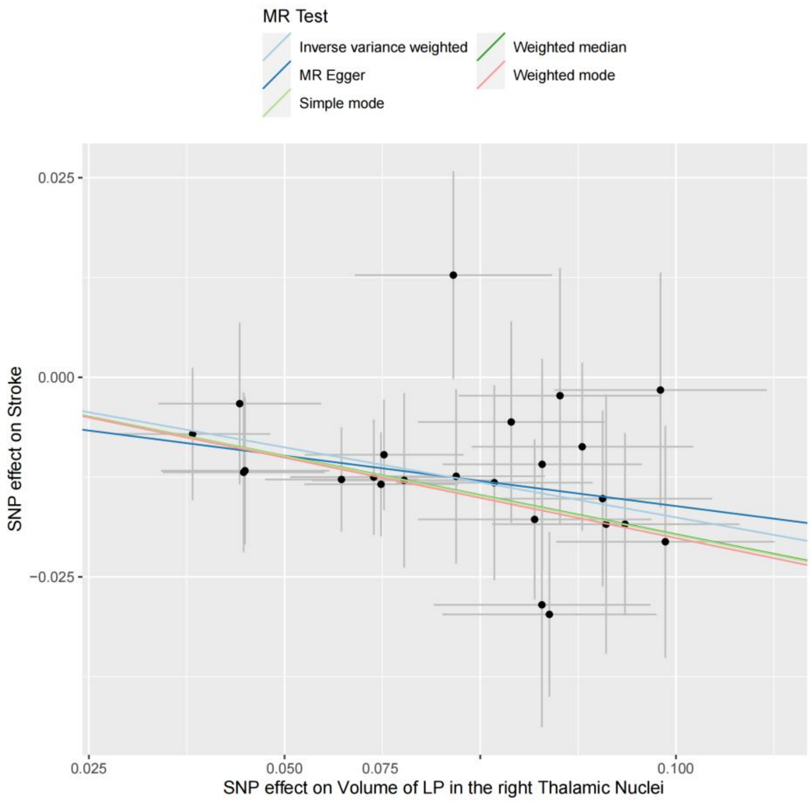


**Supplementary Figure 2** Scatter plot of individual SNP effects and estimates from different MR methods, Volume of LP in the right Thalamic Nuclei to Stroke

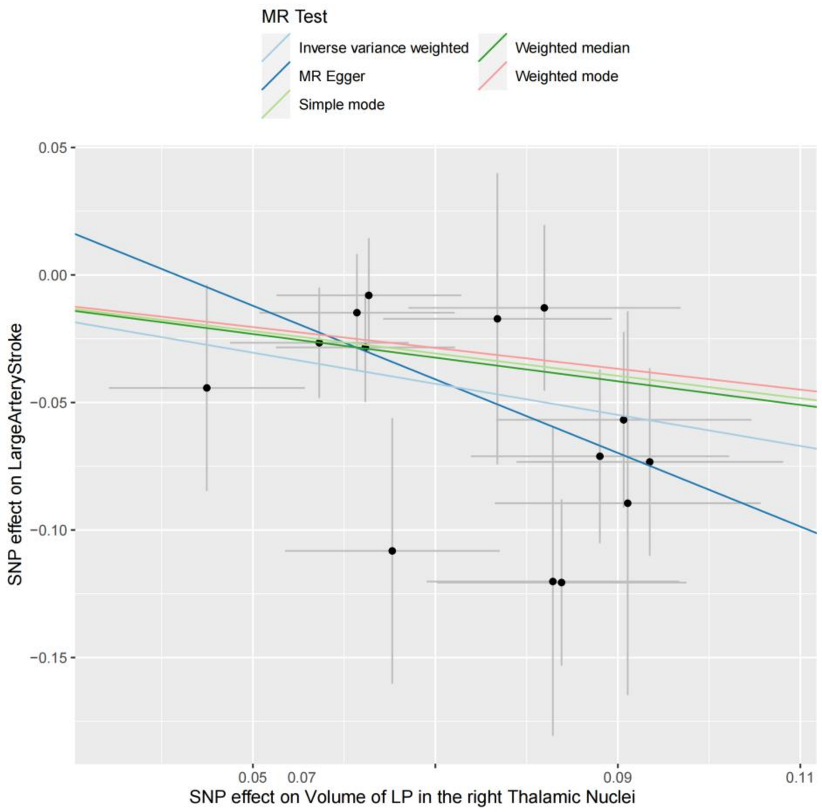


**Supplementary Figure 3** Scatter plot of individual SNP effects and estimates from different MR methods, Volume of LP in the right Thalamic Nuclei to Large Artery Stroke

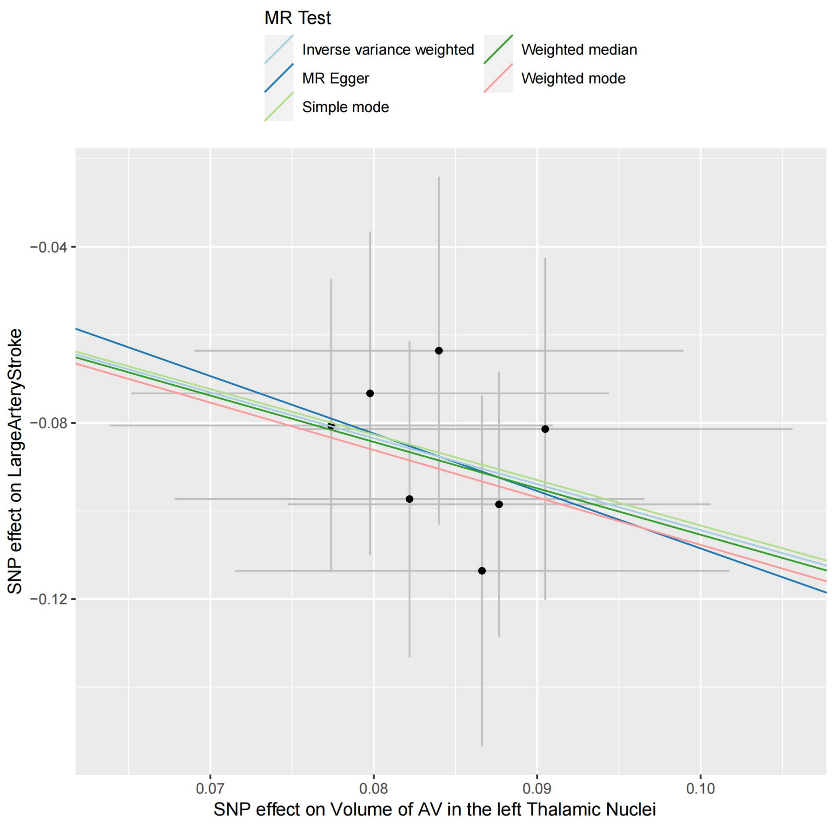


**Supplementary Figure 4** Scatter plot of individual SNP effects and estimates from different MR methods, Volume of AV in the left Thalamic Nuclei to Large Artery Stroke

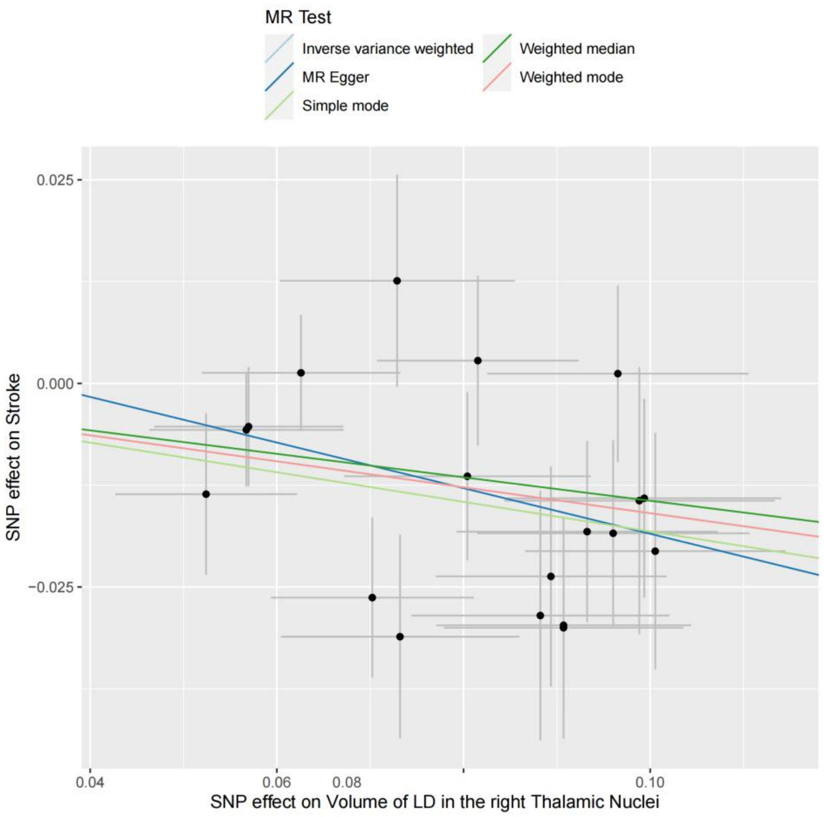


**Supplementary Figure 5** Scatter plot of individual SNP effects and estimates from different MR methods, Volume of LD in the right Thalamic Nuclei to Stroke

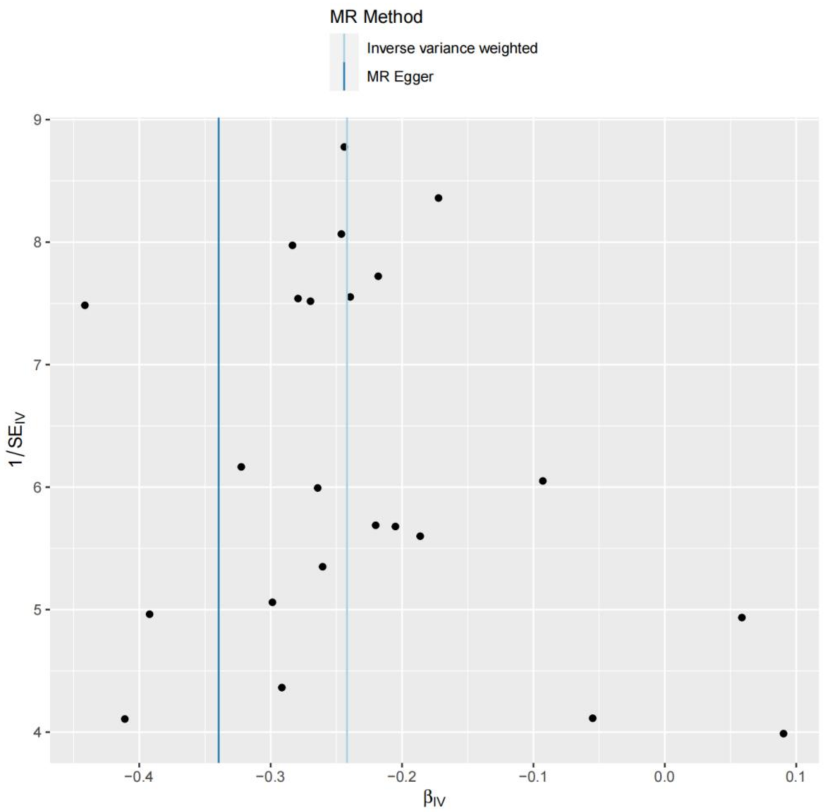


**Supplementary Figure 6** Funnel plot of Volume of LP in the right Thalamic Nuclei to Ischemic Stroke

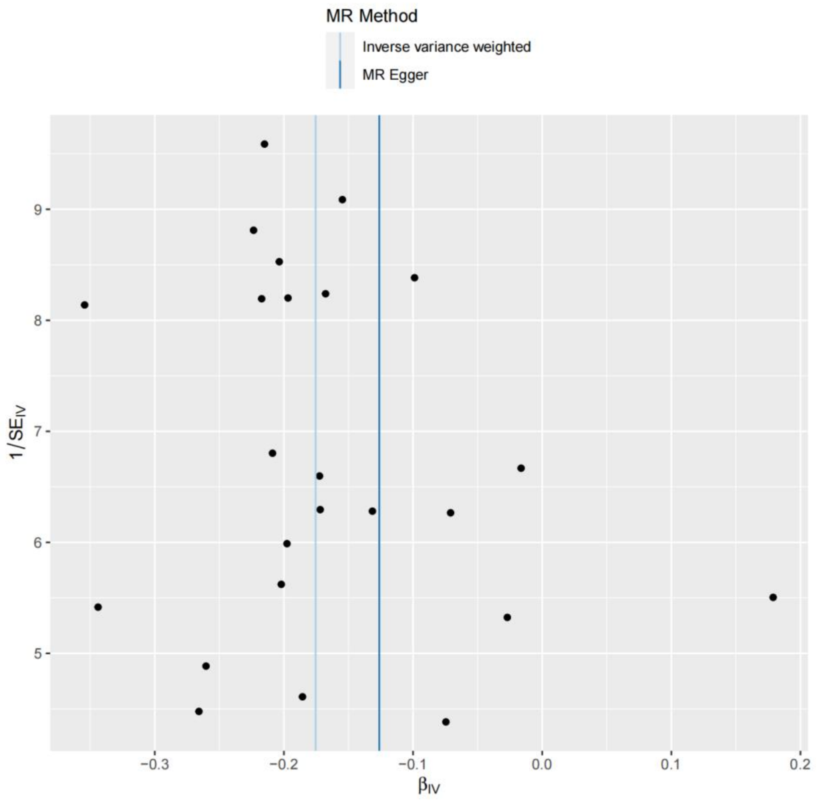


**Supplementary Figure 7** Funnel plot of Volume of LP in the right Thalamic Nuclei to Stroke

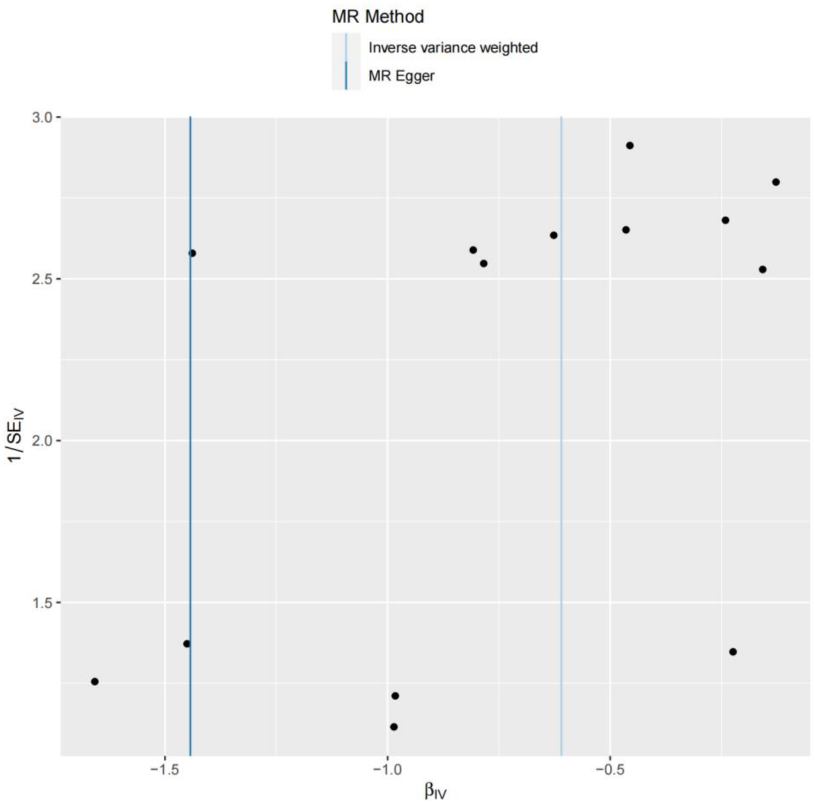


**Supplementary Figure 8** Funnel plot of Volume of LP in the right Thalamic Nuclei to Large Artery Stroke

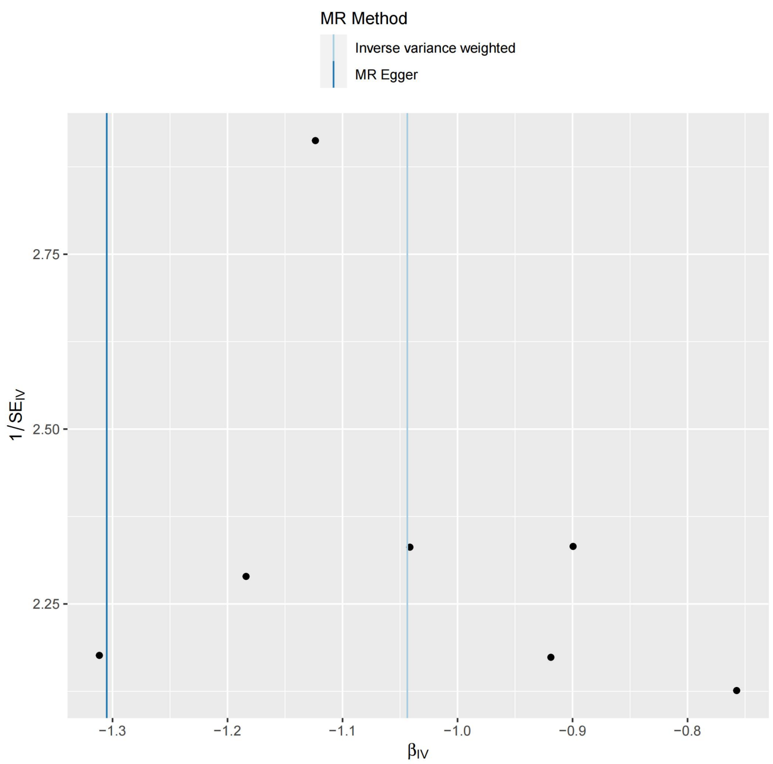


**Supplementary Figure 9** Funnel plot of Volume of AV in the left Thalamic Nuclei to Large Artery Stroke

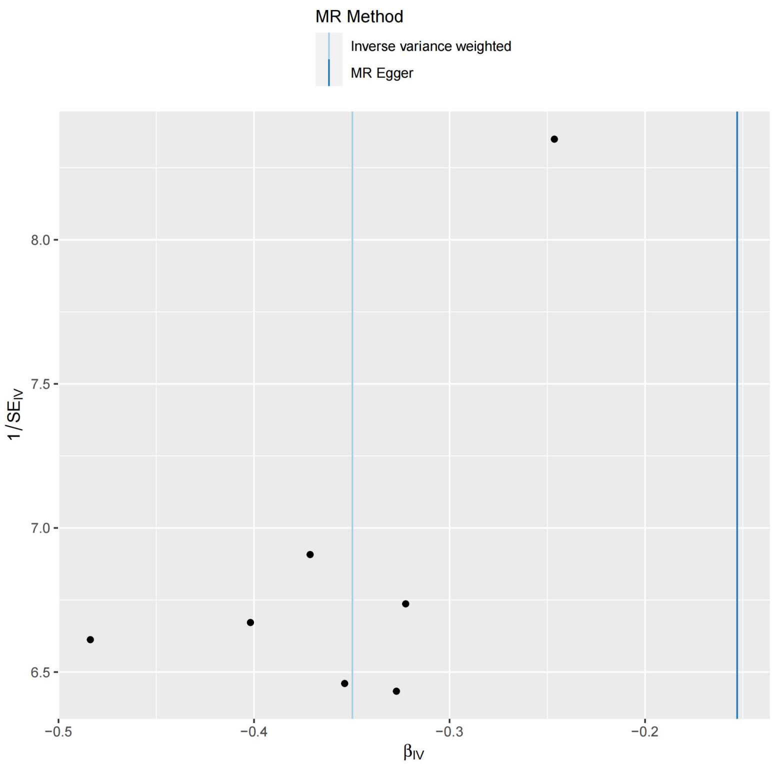


**Supplementary Figure 10** Funnel plot of Volume of AV in the left Thalamic Nuclei to Ischemic Stroke

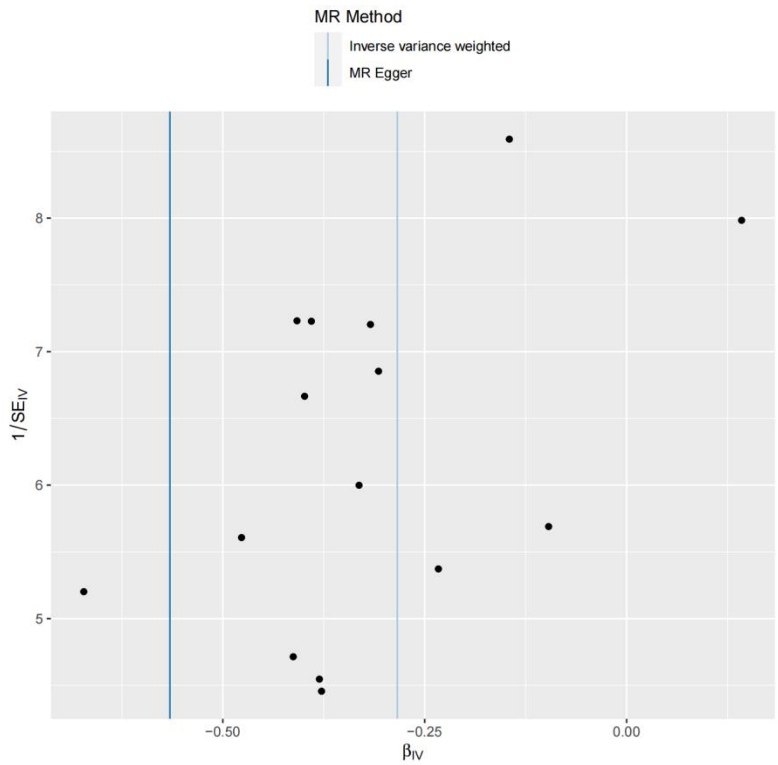


**Supplementary Figure 11** Funnel plot of Volume of CL in the right Thalamic Nuclei to Ischemic Stroke

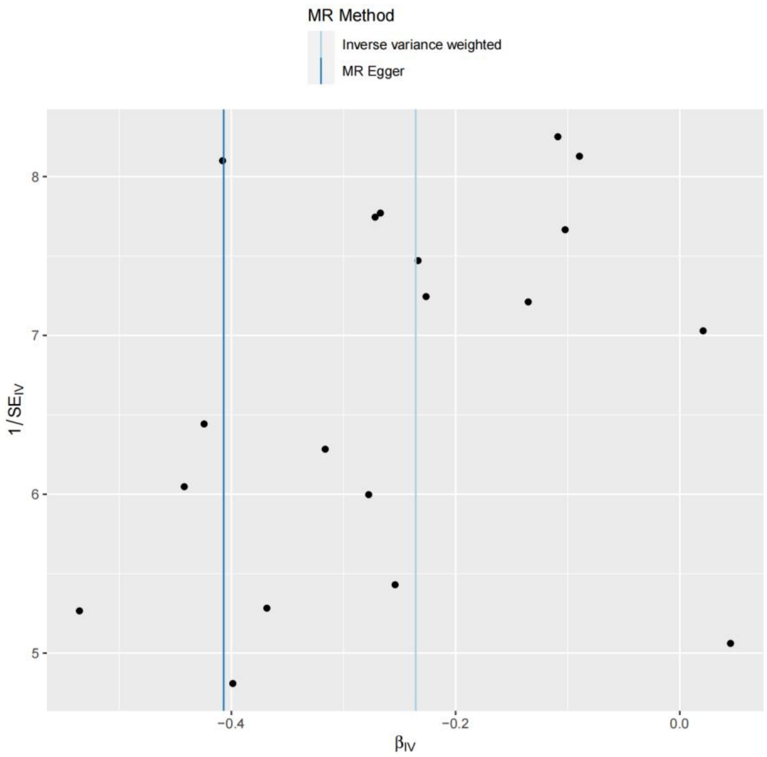


**Supplementary Figure 12** Funnel plot of Volume of LD in the right Thalamic Nuclei to Ischemic Stroke

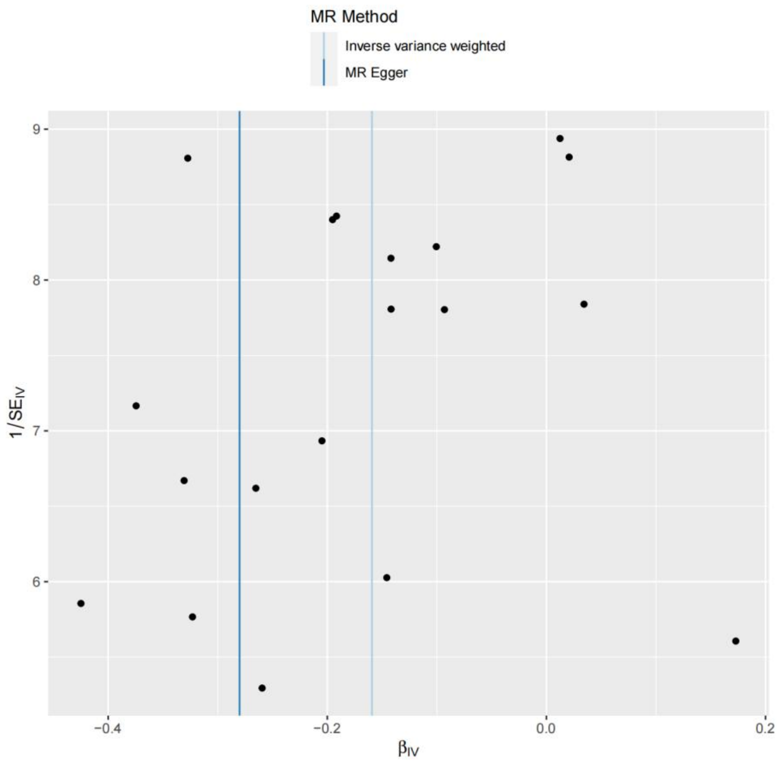


**Supplementary Figure 13** Funnel plot of Volume of LD in the right Thalamic Nuclei to Stroke

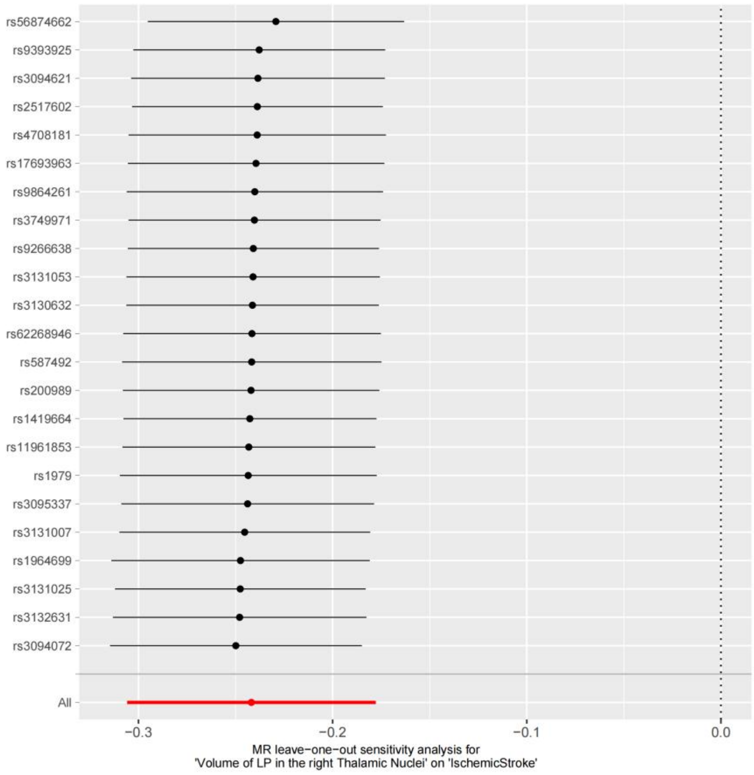


**Supplementary Figure 14** MR leave-one-out sensitivity analysis of Volume of LP in the right Thalamic Nuclei to Ischemic Stroke

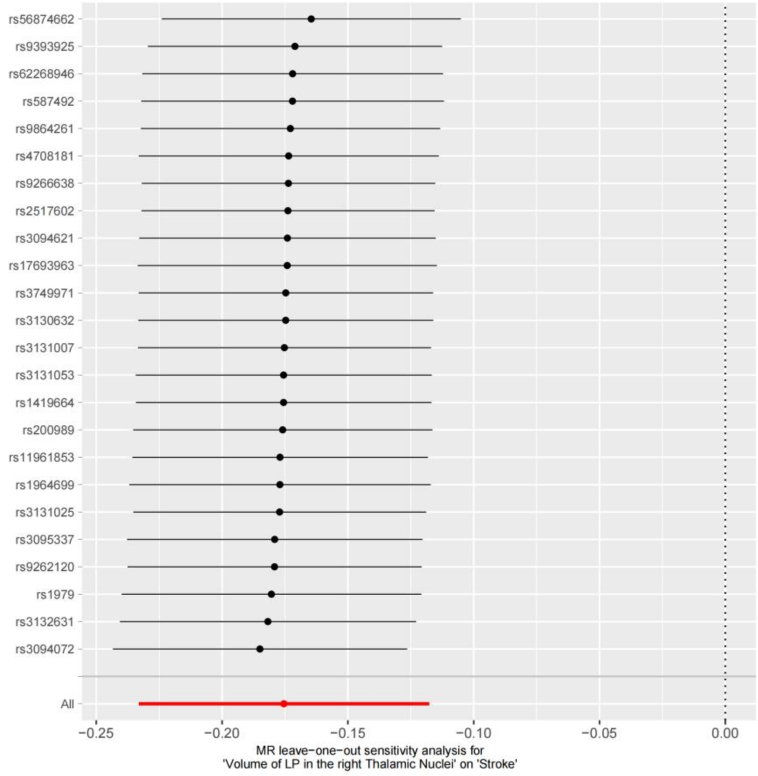


**Supplementary Figure 15** MR leave-one-out sensitivity analysis of Volume of LP in the right Thalamic Nuclei to Stroke

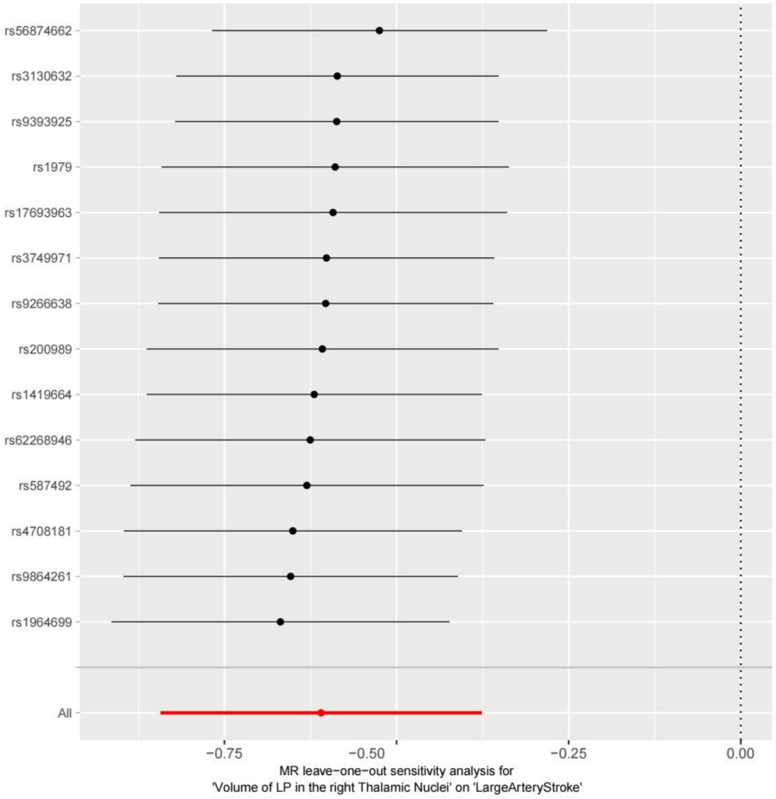


**Supplementary Figure 16** MR leave-one-out sensitivity analysis of Volume of LP in the right Thalamic Nuclei to Large Artery Stroke

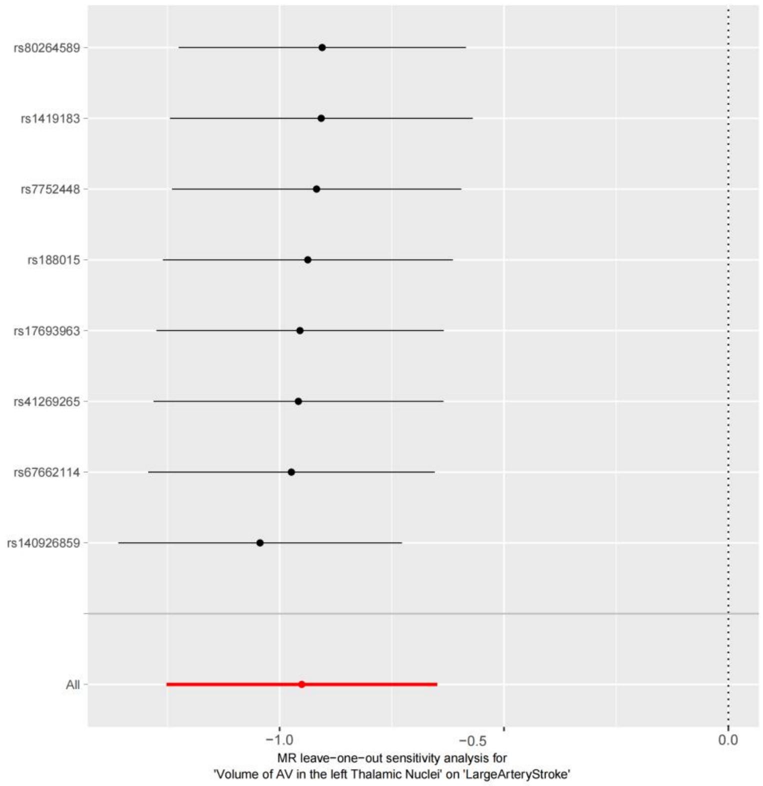


**Supplementary Figure 17** MR leave-one-out sensitivity analysis of Volume of AV in the left Thalamic Nuclei to Large Artery Stroke

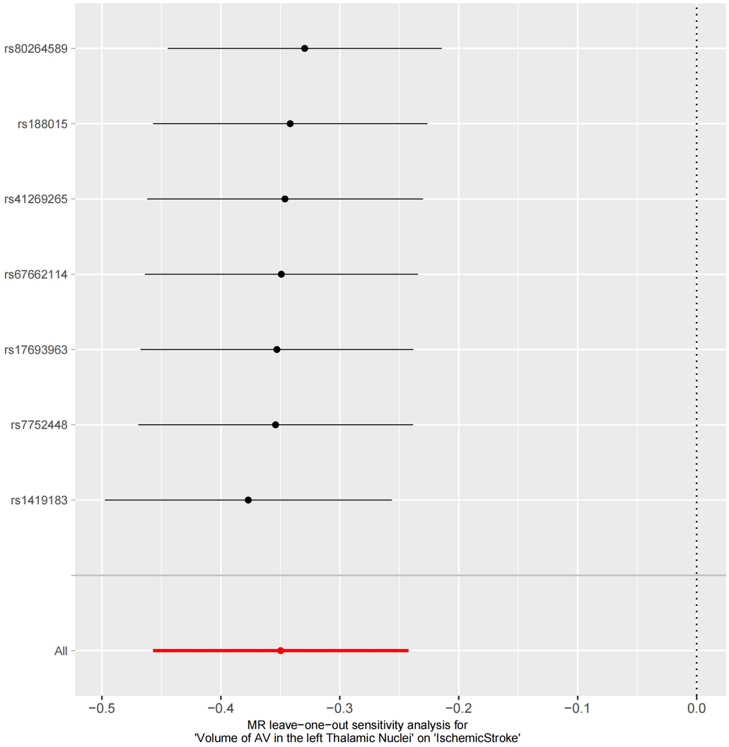


**Supplementary Figure 18** MR leave-one-out sensitivity analysis of Volume of AV in the left Thalamic Nuclei to Ischemic Stroke

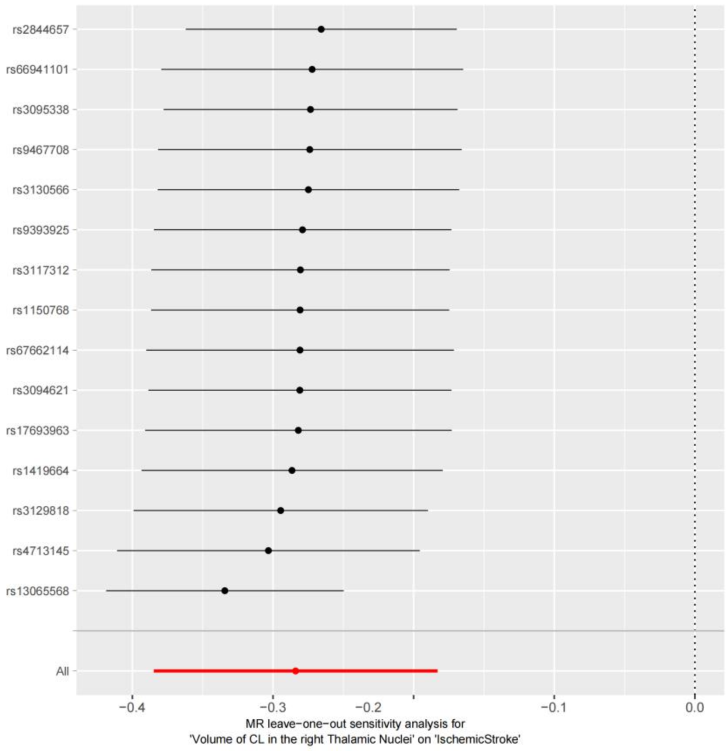


**Supplementary Figure 19** MR leave-one-out sensitivity analysis of Volume of CL in the right Thalamic Nuclei to Ischemic Stroke

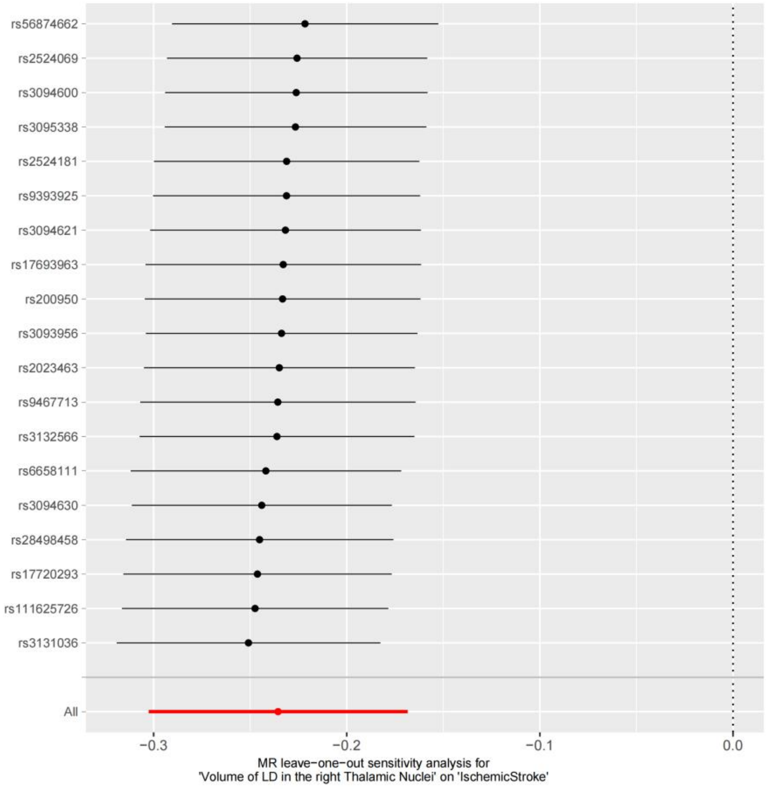


**Supplementary Figure 20** MR leave-one-out sensitivity analysis of Volume of LD in the right Thalamic Nuclei to Ischemic Stroke

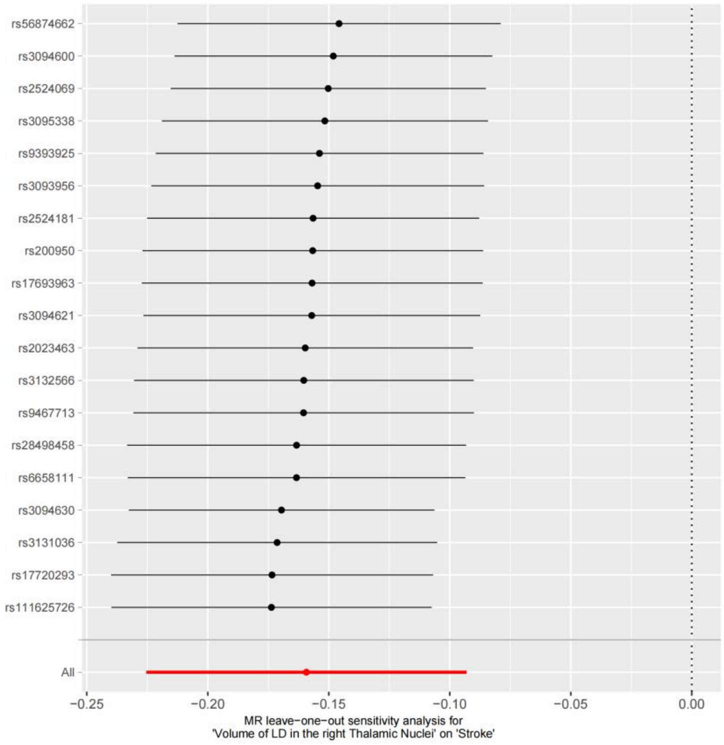


**Supplementary Figure 21** MR leave-one-out sensitivity analysis of Volume of LD in the right Thalamic Nuclei to Stroke
